## Supplementary Table 1 for "Ct threshold values, a proxy for viral load in community SARS-CoV-2 cases, demonstrate wide variation across populations and over time"

**Supplementary File 1 Month of recruitment into the COVID-19 Infection Survey**

| Month | Participants recruited | % of total 440,479 |
| --- | --- | --- |
| April 2020 | 2,206 | 0.5% |
| May 2020 | 19,652 | 4.5% |
| June 2020 | 16,106 | 3.7% |
| July 2020 | 17,350 | 3.9% |
| August 2020 | 37,473* | 8.5% |
| September 2020 | 96,155* | 21.8% |
| October 2020 | 107,731* | 24.5% |
| November 2020 | 51,511 | 11.7% |
| December 2020 | 31,678 | 7.2% |
| January 2020 | 43,454 | 9.9% |
| February 2020 | 15,602 | 3.5% |
| March 2020 | 1,561 | 0.4% |

* recruitment was increased from the middle of August 2020 to target approximately 150,000 individuals being swabbed every fortnight by the middle of October 2020 in anticipation of a second wave of infections in the autumn of 2020.

Note: recruitment started 26 April 2020 in England, 29 June 2020 in Wales, 29 July 2020 in Northern Ireland and 21 September 2020 in Scotland.
