## Supplementary Table 2 for "Ct threshold values, a proxy for viral load in community SARS-CoV-2 cases, demonstrate wide variation across populations and over time"

**Supplementary File 2: Association between characteristics and Ct values**

**Supplementary File 2A: Univariable effects and main model considering all factors**

| **Factor** | **Levels** | **n (N=27,879)** | **Median (IQR) Ct per sample unadjusted** | **p (vs reference)** | **Adjusted difference in median Ct vs reference† (95% CI)** | **p (vs reference)** |
| --- | --- | --- | --- | --- | --- | --- |
| **Independent predictors** |  |  |  |  |  |  |
| Overall (at reference category) | | **-** | **-** | **-** | 33.8 (33.6 to 34.1) |  |
| Number of genes detected | 1 | 6550 | 33.8 (32.9-34.7) | - | - | - |
|  | 2 | 1145 | 32.3 (30.9-33.4) | <0.0001 | -1.8 (-2.3 to -1.2) | <0.0001 |
|  | 2: ORF1ab+N 16nov onwards | 9867 | 26.4 (19.4-31.1) | <0.0001 | -6.9 (-7.2 to -6.6) | <0.0001 |
|  | 3 | 10317 | 25.3 (19.8-29.5) | <0.0001 | -8.2 (-8.5 to -7.9) | <0.0001 |
| Any evidence of symptoms around the positive test* | no | 13147 | 31.2 (25.4-33.5) | - | - | - |
|  | yes (any) | 14709 | 26.7 (19.9-31.7) | <0.0001 | -2.0 (-2.2 to -1.8) | <0.0001 |
|  | yes (classic**) | 10586 | 25.7 (19.4-31.0) | <0.0001 | -2.5 (-2.8 to -2.3) | <0.0001 |
|  | yes (other only) | 4123 | 29.5 (21.9-32.8) | <0.0001 | -0.9 (-1.2 to -0.7) | <0.0001 |
|  | missing | 23 | 28.1 (23.5-32.5) |  | (excluded) |  |
| Any prior positive in the study | no | 21811 | 28.4 (20.7-32.8) | - | - | - |
|  | yes | 6068 | 30.9 (26.6-32.9) | <0.0001 | +2.2 (+2.0 to +2.5) | <0.0001 |
| First test in study (ie positive at first test) | no | 24917 | 29.2 (21.9-32.8) | - | - | *-* |
|  | yes | 2962 | 29.1 (22.0-32.6) | 0.56 | +0.6 (+0.2 to +0.9) | 0.001 |
| Sex | male | 13335 | 28.9 (21.5-32.7) | - | - | - |
|  | female | 14544 | 29.5 (22.2-32.9) | <0.0001 | +0.3 (+0.1 to +0.5) | 0.001 |
| Ethnicity | white | 24902 | 29.3 (22.0-32.8) | - | - | - |
|  | non-white | 2977 | 28.6 (21.4-32.5) | <0.0001 | -0.3 (-0.6 to +0.0) | 0.08 |
| **Collinear factors with similar effects to those above** | | |  |  |  |  |
| Any evidence of symptoms at the positive test | no | 17364 | 31.0 (24.8-33.4) | - | - | - |
|  | yes (any) | 10224 | 25.2 (19.2-30.6) | <0.0001 | -2.4 (-2.6 to -2.2) | <0.0001 |
|  | yes (classic**) | 7525 | 24.6 (19.9-32.1) | <0.0001 | -2.8 (-3.0 to -2.6) | <0.0001 |
|  | yes (other only) | 2699 | 27.6 (19.9-32.1) | <0.0001 | -1.6 (-2.0 to -1.3) | <0.0001 |
|  | missing | 291 | 28.5 (21.6-32.5) |  | (excluded) |  |
| **No evidence of independent effect†** | |  |  |  |  |  |
| Age group (years/school years (SY))‡ | 2-SY6 | 1951 | 29.3 (23.1-32.9) | 0.04 |  | 0.53 |
|  | SY7-SY11 | 2169 | 28.7 (22.1-32.4) | 0.90 |  | 0.99 |
|  | Y12-24 | 2830 | 28.7 (21.8-32.4) | - (global p<0.0001) |  | - (global p=0.33) |
|  | 25-34 | 3354 | 29.0 (22.1-32.7) | 0.38 |  | 0.68 |
|  | 35-49 | 6217 | 29.2 (21.9-32.8) | 0.03 |  | 0.15 |
|  | 50-69 | 8437 | 29.3 (21.4-32.9) | 0.02 |  | 0.74 |
|  | 70+ | 2921 | 30.2 (22.1-33.3) | <0.0001 |  | 0.82 |
| Deprivation quintile ‡‡ | 1 (most deprived) | 4110 | 28.6 (21.3-32.6) | <0.0001 |  | 0.15 |
|  | 2 | 5166 | 29.0 (21.7-32.7) | <0.0001 |  | 0.30 |
|  | 3 | 5794 | 29.2 (21.8-32.8) | 0.03 |  | 0.37 |
|  | 4 | 6402 | 29.3 (22.1-32.9) | 0.06 |  | 0.53 |
|  | 5 (least deprived) | 6407 | 29.6 (22.1-33.0) | - (global p<0.0001) |  | - (global p=0.67) |
| **Factor reflecting potential consequence** | |  |  |  |  |  |
| Anyone else in household ever positive | no | 14555 | 31.0 (24.6-33.5) | - |  | *-* |
|  | yes | 13324 | 26.8 (19.9-31.6) | <0.0001 | -1.4 (-1.6 to -1.2) | <0.0001 |

* any evidence of symptoms at the visit where the swab was positive or visits either side of this, regardless of time.

** cough, fever or loss of taste or smell.

† adjusted column shows the effect of adding this variable into a model including number of genes detected, evidence of symptoms around the test, whether the participant had any prior positive in the study, whether the positive test was the participant’s first test result in the study, sex and ethnicity (27,856 complete cases). Results similar including symptoms at the test instead (27,588 complete cases).

‡ linear effect of age on Ct p<0.0001 univariable, p=0.34 multivariable; non-linear <0.0001 (p=0.0004 for non-linearity) and 0.45 respectively.

‡‡ linear effect of deprivation score on Ct p<0.0001 univariable (no evidence of non-linearity), p=0.17 multivariable; non-linear 0.66 respectively.

Note: p-values (vs the reference category) from univariable or multivariable median regression. Main multivariable regression models include number of genes detected, evidence of symptoms around the test, whether the participant had any prior positive in the study, whether the positive test was the participant’s first test result in the study, sex and ethnicity. All multivariable associations similar adjusting for calendar date of visit using a 5 knot natural cubic spline.

**Supplementary File 2B: Multivariable model excluding potential mediators of effects of demographics**

| **Factor** | **Levels** | **Adjusted difference in median Ct vs reference† (95% CI)** | **p (vs reference)** |
| --- | --- | --- | --- |
| Overall (at reference category) | | 27.8 (27.3 to 28.3) | **-** |
| Any prior positive in the study | no (ie first positive per-participant) | - | - |
|  | yes | +2.6 (+2.3 to +3.0) | <0.0001 |
| First test in study (i.e. positive at first study test) | no | - | - |
|  | yes | +1.0 (+0.5 to +1.4) | <0.0001 |
| Sex | male | - | - |
|  | female | +0.6 (+0.3 to +0.9) | <0.0001 |
| Ethnicity | white | - | - |
|  | non-white | -0.3 (-0.8 to +0.1) | 0.13 |
| Age group (years/school years (SY))‡ | 2-SY6 | +1.0 (+0.3 to +1.7) | 0.004 |
|  | SY7-SY11 | +0.2 (-0.5 to +0.8) | 0.62 |
|  | Y12-24 | - | - (global p=0.0001) |
|  | 25-34 | +0.5 (-0.0 to +1.1) | 0.07 |
|  | 35-49 | +0.5 (+0.0 to +1.0) | 0.05 |
|  | 50-69 | +0.6 (+0.1 to +1.1) | 0.02 |
|  | 70+ | +1.4 (+0.8 to +2.0) | <0.0001 |
| Deprivation quintile ‡‡ | 1 (most deprived) | -1.0 (-1.5 to -0.6) | <0.0001 |
|  | 2 | -0.6 (-1.0 to -0.2) | 0.005 |
|  | 3 | -0.4 (-0.8 to -0.0) | 0.05 |
|  | 4 | -0.4 (-0.8 to -0.0) | 0.03 |
|  | 5 (least deprived) | - | - (global p=0.0005) |

‡ non-linear effect of age on Ct p<0.0001.

‡‡ linear effect of deprivation score on Ct p<0.0001 (+0.2 (+0.1 to +0.3) higher Ct per quintile higher; non-linear p=0.13.

Results similar adjusted for calendar date using 5 knot natural cubic spline.
