## Supplementary figures and images for "Ct threshold values, a proxy for viral load in community SARS-CoV-2 cases, demonstrate wide variation across populations and over time"

### Supplementary Figure 1

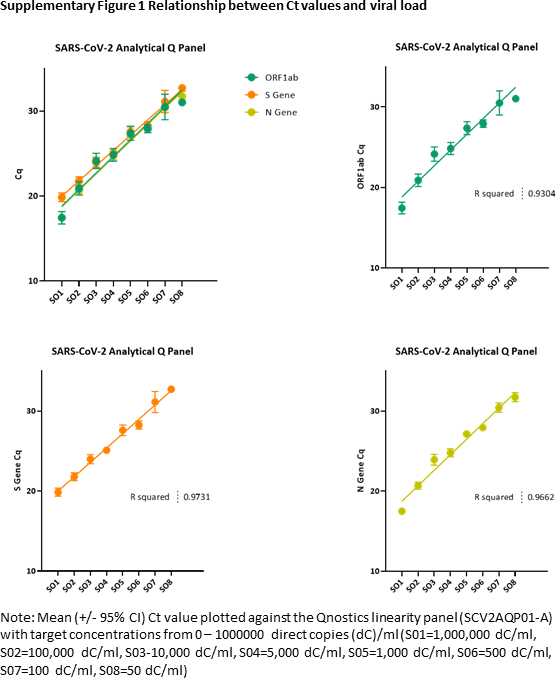

### Supplementary Figure 2

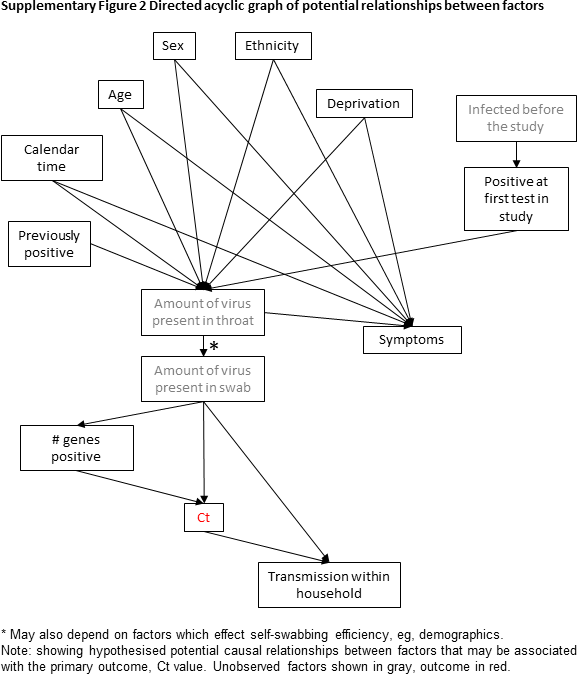
